# Changes in vaginal *Ureaplasma* and *Lactobacillus* due to antibiotic regimen for premature rupture of membranes

**DOI:** 10.1101/2024.06.28.24309657

**Authors:** Haruna Kawaguchi, Yukiko Nakura, Ryo Yamamoto, Shusaku Hayashi, Makoto Takeuchi, Keisuke Ishii, Itaru Yanagihara

**Affiliations:** Department of Maternal Fetal Medicine, Osaka Women’s and Children’s Hospital, Osaka, Japan; Department of Developmental Medicine, Research Institute, Osaka Women’s and Children’s Hospital, Osaka, Japan; Department of Pathology, Osaka Women’s and Children’s Hospital, Osaka, Japan

**Keywords:** Azithromycin, bronchopulmonary dysplasia, funisitis, histological chorioamnionitis, neonatal infection

## Abstract

Preterm premature rupture of membranes (PPROM) is associated with preterm delivery and neonatal complications. PPROM is often complicated by intra-amniotic inflammation and/or microbial invasion of the amniotic cavity with *Ureaplasma* or *Mycoplasma*. Various prophylactic antibiotic therapies have been proposed to prolong latency between PPROM and delivery, reduce the risk of clinical chorioamnionitis, and improve neonatal complications. However, information on the potential of azithromycin administration to reduce the microbial load of vaginal *Ureaplasma* and *Mycoplasma* remains lacking. This prospective cohort study included singleton pregnancies managed with prophylactic antibiotics for PPROM at less than 36 weeks of gestation. All patients received the standard antibiotic regimen for PPROM, which consisted of a single oral azithromycin and intravenous ampicillin every for 2 days followed by 5 days of oral amoxicillin. Vaginal swabs samples were collected when PPROM was confirmed and after the antibiotic regimen administration. The main outcome measures were to investigate the changes in vaginal *Ureaplasma, Mycoplasma,* and *Lactobacillus spp.* due to the antibiotic regimen. In addition, the association between the presence and changes in vaginal *Ureaplasma* and *Mycoplasma*, pregnancy outcomes, and neonatal complications were examined. Out of 82 eligible PPROM, 51 had positive vaginal *Ureaplasma*. Thirty-six patients (52.2%) completed the antibiotic regimen. Among those with positive vaginal *Ureaplasma* who completed the antibiotic regimen, 75% experienced an increase in vaginal *Ureaplasma* levels. For those who delivered before completing all antibiotic doses, 40% had increased vaginal *Ureaplasma* levels. Furthermore, the antibiotic regimen resulted in decreased *Lactobacillus spp.* in almost all cases. However, vaginal *Ureaplasma* changes were not found to be associated with neonatal sepsis or bronchopulmonary dysplasia. This suggests that *Ureaplasma* became resistant to azithromycin. Future studies are needed to revalidate current antibiotic therapy for PPROM.

## Introduction

Preterm premature rupture of membranes (PPROM) occurs in 2%−3% of pregnancies and is associated with preterm delivery, neonatal complications, and maternal sepsis.(1–4) PPROM is often complicated by intra-amniotic inflammation and/or microbial invasion of the amniotic cavity with *Ureaplasma* or *Mycoplasma*. (5) Both are linked to neonatal infections and chronic lung disease. (6) Various prophylactic antibiotic regimens were proposed to prolong the latency between PPROM and delivery, reduce the risk of clinical chorioamnionitis, and improve neonatal outcomes. (7–10) The American College of Obstetricians and Gynecologists recommends using erythromycin and ampicillin, but a regimen with azithromycin has been suggested as an alternative to erythromycin. (8, 11, 12) At our institution, patients with PPROM receive a standard antibiotic regimen consisting of a single oral dose of azithromycin and intravenous ampicillin for 2 days followed by 5 days of oral amoxicillin. However, it is unclear whether administering azithromycin can reduce the microbial load of vaginal *Ureaplasma* and *Mycoplasma* DNA. Therefore, this study aimed to investigate the changes in vaginal *Ureaplasma*, *Mycoplasma*, and *Lactobacillus*, as well as maternal blood *Ureaplasma*, resulting from the antibiotic regimen used for PPROM cases. In addition, the association between the presence and changes in vaginal *Ureaplasma* and *Mycoplasma,* pregnancy outcomes, and neonatal complications were examined, and that aim was achieved.

## Materials and Methods

This single-center, prospective cohort study included singleton pregnancies managed with prophylactic antibiotics for PPROM at less than 36 weeks of gestation between October 2019 and December 2021. Patients under the age of 20, those who received other antibiotics within 2 weeks of PPROM, and those allergic to ampicillin and azithromycin were excluded. All relevant national regulations and institutional policies were compiled, and the study was conducted under the tenets of the Helsinki Declaration. It was approved by the ethical review board of Osaka Women’s and Children’s Hospital (approval number, 1211; date of approval, June 3, 2019). Written informed consent was obtained from all study subjects. Recruitment of subjects for this study began on October 1, 2019 and ended on December 31, 2021. The standard deviation of the amount of vaginal *Ureaplasma* colonization was 10^2.7^. To have a 90% power to detect changes above 10^2^ colony-forming units/mL, the study aimed to enroll 80 patients.

The perinatal management protocol for PPROM at our institution is as follows. The diagnosis of PPROM is confirmed by observing amniotic fluid leakage from the cervix or pooling in the vagina, along with positive diagnostic tests measuring vaginal pH. In cases where the diagnosis is not clearly confirmed, an immunochromatographic strip test with insulin-like growth factor-binding protein-1 (ActimProma^ℝ^) is used to detect amniotic fluid in the vagina or ultrasound evaluation is used to confirm the presence of oligohydramnios. All patients were admitted to the hospital and received the standard antibiotic regimen for PPROM before 36 weeks of gestation with intravenous ampicillin (2 g dose every 6 hours) for 48 hours and single oral azithromycin (1 g dose) followed by oral amoxicillin (500 mg dose every 8 hours) for 5 days. Betamethasone was administrated for patients with gestational age less than 34 weeks and imminent delivery. Intravenous magnesium sulfate and/or ritodrine hydrochloride and/or oral nifedipine as tocolytic agents were administered when patients have short cervical length and frequent uterine contraction. Indications for delivery included clinical chorioamnionitis, onset of labor, nonreassuring fetal status, or gestational age beyond 36 weeks. Clinical chorioamnionitis was diagnosed based on the presence of maternal fever (≥38.0°C) and two or more clinical signs: maternal leukocytosis (white blood cell count >15,000/mm^3^), maternal tachycardia (heart rate > 100 beats/min), fetal tachycardia (heart rate > 160 beats/min), uterine tenderness, and purulent or foul-smelling amniotic fluid or vaginal discharge. (13)

Vaginal swabs and maternal blood samples were collected from women when the diagnosis of PPROM was confirmed. The quantity of vaginal secretions was measured by weighing the swab tube before and after the collection. Vaginal swabs were taken from the lateral vaginal walls and the anterior and posterior fornix, and they were transferred to a cobas® polymerase chain reaction (PCR) Media Dual Swab Sample Kit from Roche Diagnostics GmbH, Germany. Vaginal swabs and maternal blood samples were retaken after the completion of the antibiotic regimen. Scrapings of the placental surface were also performed. If delivery occurred before the completion of the regimen, vaginal swabs and maternal blood samples were collected before the delivery. These results were not available to managing clinicians. The amount and species of *Ureaplasma* collected from the vaginal secretions, along with the amount of *Lactobacillus* spp. and *Mycoplasma (M.) hominis* and the presence of inerolysin (pore-forming toxin produced by *Lactobacillus (L.) iners* that can cause preterm labor) were determined using real-time PCR. The amount of *Ureaplasma* was also measured from maternal blood and placental surface. An increase or decrease of 10^2^ or more in DNA copy numbers was considered significant.

This study investigated neonatal and infant mortality and any of the following adverse perinatal outcomes for up to 6 months of age: neonatal intensive care unit admission, mechanical ventilation, respiratory distress syndrome, retinopathy of prematurity, intraventricular hemorrhage 3 or 4 grade, periventricular leukomalacia, necrotizing enterocolitis, neonatal sepsis, bronchopulmonary dysplasia (BPD), and congenital anomaly. BPD is a chronic lung disease, which is most commonly seen in premature infants requiring respiratory support, including oxygen supplementation at 36 weeks corrected gestational age. (14) Neonatal sepsis is defined as the presence of a positive blood culture. (15)

In this study, changes in vaginal *Ureaplasma, Mycoplasma*, and *Lactobacillus* are divided into two groups: complete and incomplete antibiotic protocols. Cases of complete protocol are defined as cases in which all antibiotic regimens had been completed. Cases of incomplete protocol are those who delivered in the middle of their antibiotic regimen, but all of them had received at least one intravenous ampicillin and oral azithromycin administration.

### DNA extraction and quantitative real-time PCR of *Ureaplasma*

DNA was extracted from vaginal swab and maternal blood samples using a Maxwell® RSC Blood DNA Kit (AS1400, Promega, Japan). The DNA copy number of *Ureaplasma* was measured using QuantStudio 5Real-Time PCR System (Thermo Fisher Scientific) targeted for the 16S ribosomal RNA gene. The amplification reaction mixtures comprised 5 mL of TaqMan Fast Advanced Master Mix (Thermo Fisher Scientific), primers (0.9 μM each), probe (Thermo Fisher Scientific, 0.25 μM), and 1 μL of DNA sample; this was adjusted to 10 μL with DDW. The primers and probe used were as follows: 16S-Urea F1(AGGCATGCGTCTAGGGTAGGA), 16S-Urea R1(ACGTTCTCGTAGGGATACCTTGTTA), and 16S-Urea-FAM-MGB probe (FAM-CGGTGACTGGAGTTAA-MGB). The PCR conditions were as follows: predenaturation at 95°C for 20 s, 40 cycles of denaturation at 95°C for 1 s, and annealing at 58 C for 20 s. The standard curve of *Ureaplasma* was determined using serial dilutions with copy numbers of 1.0 × 10^1^ to 1.0 × 10^6^ copies per reaction mixture by real-time PCR. In each run with clinical specimens, negative control and standards were included. To differentiate *Ureaplasma* (*U.) parvum* from *U. urealyticum*, the following primers and probes were used: ureG F2(CAACATTTAGTCCAGATTTAG), ureG R2(TAGCACCAACATAAGGAG), Up-FAM-MGB probe (FAM-TTGACCACCCTTACGAG-MGB), and Uu-VIC-MGB probe (VIC-TTGTCCGCCTTTACGAG-MGB). The amplification reaction mixtures comprised 5 mL of TaqMan Fast Advanced Master Mix (Thermo Fisher Scientific), primers (0.9 μM each), probes (Thermo Fisher Scientific, 0.2 μM each), and 1 μL of sample DNA; this was adjusted to 10 μL with DDW.

### Quantitative real-time PCR analysis of *M. hominis*

The primers and probe used were as follows: Mh-yidC-F(ACCCGGTTTAGTGAGTTTGCT), Mh-yidC-R (CCTCAGTTTATTGCATTGCCA), and Mh-yidC-VIC probe (VIC-AACAAGCAACCTGATATT-MGB). The PCR conditions were as follows: predenaturation at 95°C for 20 s, 35 cycles of denaturation at 95°C for 1 s, and annealing at 60°C for 20 s.

Quantitative real-time PCR of *Lactobacillus* species and inerolysin detection The primers for *Lactobacillus* spp. used were Lacto-F (TGGAAACAGRTGCTAATACCG) and Lacto-R (GTCCATTGTGGAAGATTCCC).(16) The primers for inerolysin were; inerolysin-F(CGTGGTCGTACATCAGGCTT) and inerolysin-R(TTCTCCACCATTCCCATGCC). PCRs were performed in a reaction volume of 10 μL, each containing 0.5 µM of inerolysin-F and inerolysin-R primers, 5 μL of PowerUP^TM^ SYBR^TM^ Green Master Mix (Thermo Fisher Scientific), and 1.0 μL of genomic DNA. PCR amplification was performed under the following conditions: initial cycle of 95°C for 2 min, 35 cycles of 95°C for 1 s, 58°C for 15 s, and 72°C for 60 s.

### 2.4 Statistical analysis

Categorical and continuous variables were analyzed using chi-square or Fisher’s exact tests and compared using Kruskal–Wallis test. Multivariate logistic regression analyses were performed to examine the adjusted odds ratios (aOR) and 95% confidence intervals (CI) for changes in *Ureaplasma* and perinatal outcomes. Statistical significance was set at P <0.05. Two-sided P-values were reported, and analyses were conducted using a JMP software (version 14; SAS Institute, Cary, NC, USA).

## Results

In total, 82 patients with PPROM were included, and 51 were positive for vaginal *Ureaplasma.* Thirty-six patients (52.2%) completed the standard antibiotic regimen. (Figure 1) Of those who had positive vaginal *Ureaplasma* and completed the standard antibiotic regimen, vaginal *Ureaplasma* increased in 18 patients (75%). Among patients with positive vaginal *Ureaplasma* who delivered before the completion of all antibiotic doses, vaginal *Ureaplasma* increased in eight patients (40%).

**Fig 1.**
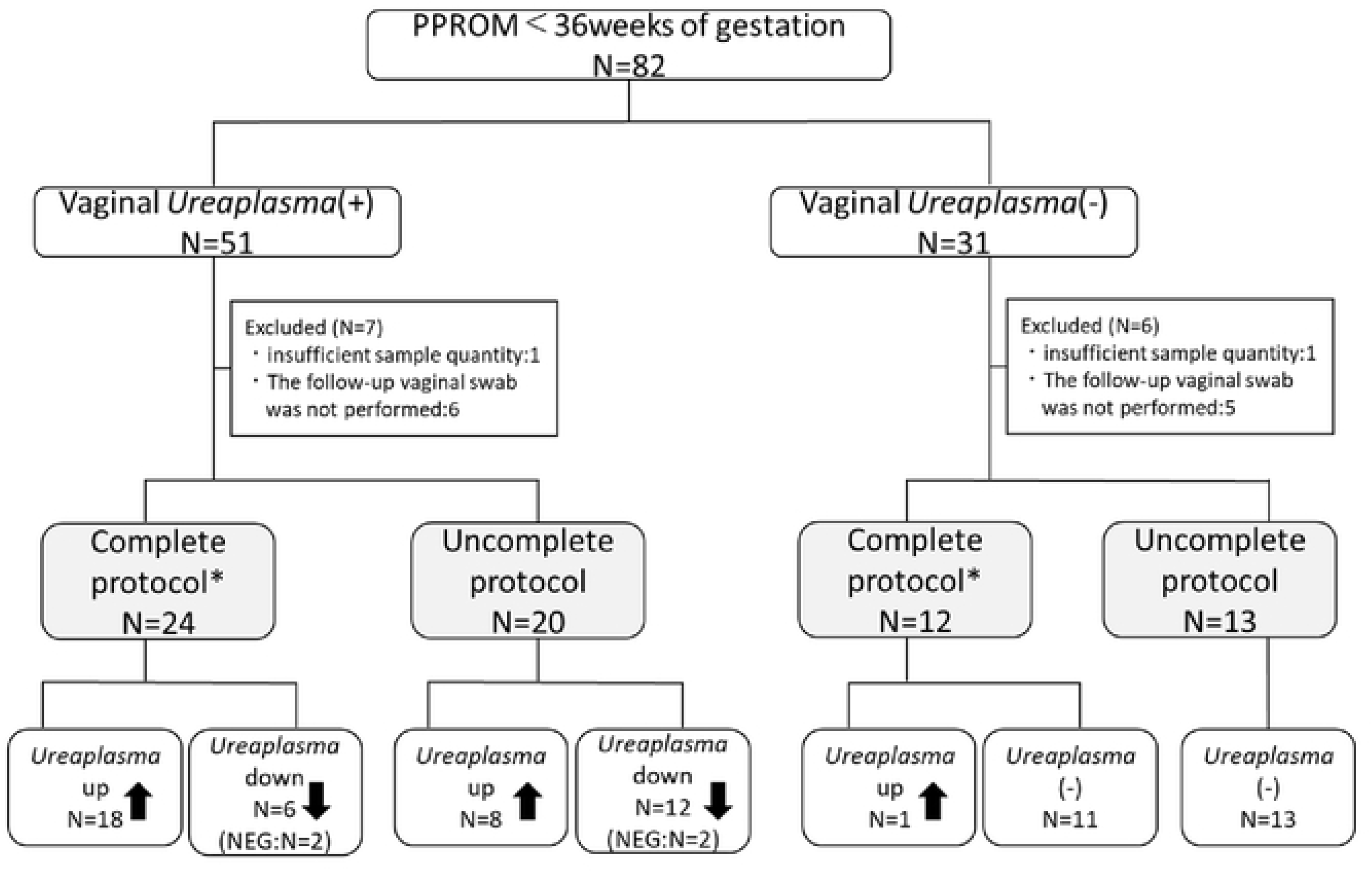
Flowchart of the study population. A total of 82 patients with PPROM were eligible for this study. Vaginal *Ureaplasma* was positive in 51 patients and negative in 31 patients. Of the patients who had positive vaginal *Ureaplasma* and completed the standard antibiotic regimen, 18 (75%) had increased vaginal *Ureaplasma* after treatment. Among patients with a positive vaginal *Ureaplasma* delivered before completion of all antibiotic doses, vaginal *Ureaplasma* increased in eight patients (40%). PPROM, preterm premature rupture of membranes; NEG, negative * intravenous 2 g ampicillin every 6 hours for 2 days, a single oral dose of 1 g azithromycin, and 5 days of oral 500 mg amoxicillin every 8 hours.

Table 1 shows the maternal characteristics and perinatal findings classified into three groups based on vaginal *Ureaplasma* changes: *Ureaplasma*-negative before and after the antibiotic regimen, *Ureaplasma*-positive that decreased after the antibiotic regimen, and *Ureaplasma*-positive that increased after the antibiotic regimen.

**Table 1.** Maternal characteristics and perinatal findings, classified into three groups based on vaginal *Ureaplasma* changes by antibiotic treatment.

|  |  | *Ureaplasma*<br>(-) at the time<br>of PPROM | *Ureaplasma* (+) at the time<br>of PPROM |  |  |
| --- | --- | --- | --- | --- | --- |
|  | Total |  | Decreased<br>after<br>treatment | Increased<br>after<br>treatment | *p* |
|  | N = 69 | N = 24 | N = 18 | N = 27 |  |
| maternal age at<br>delivery | 33(23–46) | 35 (26–46) | 31.5 (22–41) | 33 (23–42) | 0.21 |
| nulliparous | 40 | 10 (41.7) | 14 (77.8) | 16 (59.3) | 0.06 |
| history of<br>preterm birth | 4 | 1 (4.2) | 0 | 3 (11.1) | 0.27 |
| Obesity (BMI<br>≥25) | 11 | 4 (16.7) | 4 (22.2) | 3 (11.1) | 0.6 |
| ART | 10 | 3 (12.5) | 2 (11.1) | 5 (18.5) | 0.74 |
| tobacco use<br>during<br>pregnancy | 4 | 1 (4.2) | 1 (5.6) | 2 (7.4) | 0.88 |
| CHT | 1 | 1 (4.2) | 0 | 0 | - |
| DM | 0 | 0 | 0 | 0 | - |
| history of<br>cervical<br>conization | 7 | 1 (4.2) | 0 | 6 (22.2) | 0.02<br>6 |
| cerclage | 6 | 2 (8.3) | 2 (11.1) | 2 (7.4) | 0.91 |
| antenatal<br>corticosteroid | 38 | 14 (58.3) | 10 (55.6) | 14 (51.9) | 0.9 |
| tocolysis<sup>a</sup> | 39 | 13 (54.2) | 10 (55.6) | 16 (59.3) | 0.93 |
| HDP | 4 | 2 (8.3) | 0 | 2 (7.4) | 0.47 |
| GDM | 7 | 1 (4.6) | 3 (17.6) | 3 (11.5) | 0.42 |
| antibiotics | 69 | 24 (100) | 18 (100) | 27 (100) | - |
| complete<br>protocol<sup>b</sup> | 36 | 11 (45.8) | 6 (33.3) | 19 (70.4) | 0.03<br>8 |
| azithromycin | 69 | 24 (100) | 18 (100) | 27 (100) | - |
| GA at PPROM | 31 + 6 (12 +<br>5~35 + 6) | 32 + 3 (20 + 0<br>~35 + 6) | 32 + 2 (17 +<br>6~35 + 3) | 31 + 3 (12<br>+ 5~35 +<br>2) | 0.43 |
| GA at delivery | 33 + 1 (17 +<br>5~36 + 4) | 33 + 2 (23 + 0<br>~36 + 4) | 33 + 3 (18 +<br>1~36 + 1) | 32 + 4 (17<br>+ 5~36 +<br>3) | 0.83 |
| Latency<sup>c</sup> | 7 (0–86) | 5 (0–86) | 3.5 (0–21) | 10 (2–52) | 0.01<br>2 |
| Latency < 7days | 33 | 13 (54.2) | 12 (66.7) | 8 (29.6) | 0.038 |
| CS | 25 | 10 (41.7) | 3 (16.7) | 12 (44.4) | 0.13 |
| hCAM | 39 | 12 (54.6) | 8 (50) | 19 (76) | 0.17 |
| Funisitis | 30 | 9 (40.9) | 4 (25) | 17 (68) | 0.02 |

Data are expressed as the median (range) or n (%)
<sup>a</sup> Tocolysis agents are intravenous magnesium sulfate and/or ritodrine hydrochloride
and/or oral nifedipine.
<sup>b</sup> Complete protocol is defined as the cases in which all standard antibiotic regimens
(intravenous 2 g ampicillin every 6 hours for 2 days, one oral dose of 1 g azithromycin,
and 5 days of oral 500 mg amoxicillin every 8 hours) had been completed.
<sup>c</sup> The latency is defined as the period between PPROM and delivery.
BMI, body mass index; ART, assisted reproductive technology; CHT, chronic
hypertension; DM, diabetes mellitus; HDP, hypertensive disorders of pregnancy; GDM,
gestational diabetes mellitus; GA, gestational age; PPROM, preterm premature rupture
of membranes; CS, cesarean section; hCAM, histological chorioamnionitis

The median gestational week at PPROM and delivery did not differ among the three groups. However, latency from PPROM to delivery was significantly prolonged in the group with increased *Ureaplasma* after the antibiotic regimen. There was no difference in the incidence of histological chorioamnionitis (hCAM), but funisitis was significantly more common in the group with increased *Ureaplasma* after the antibiotic regimen (*p* = 0.02).

Figure 2 shows the changes in vaginal *Ureaplasma* spp. before and after the antibiotic regimen. The median microbial load of vaginal *Ureaplasma* DNA at the time of PPROM was 1.9 × 10^7^ (1.8 × 10^2^ − 4.6 × 10^9^). Many cases had increased vaginal *Ureaplasma*, especially in patients who completed the antibiotic regimen. As shown in Figure 3, *Lactobacillus* spp. decreased after the antibiotic regimen in almost all cases. Table 2 shows the vaginal and maternal blood *Ureaplasma*, vaginal *M. hominis*, *Lactobacillus* spp., and inerolysin classified by vaginal *Ureaplasma* changes. All *M. hominis*-positive cases were positive for vaginal *Ureaplasma*. Three patients had a positive vaginal *M. hominis* result at the time of PPROM and did not decrease in all cases after antibiotic regimen, but rather turned positive in three new cases. The loss of *Lactobacillus* spp. was more common in patients who completed the antibiotic regimen, especially in the group with increased *Ureaplasma*. hCAM and funisitis were more common in those with completed antibiotic regimen, regardless of vaginal *Ureaplasma* changes due to the antibiotic regimen and presence of *Ureaplasma* at the time of PPROM (aOR,7.5; 95% CI; 2.1–26.6; *P* = 0.002, aOR4.5; 95%CI 1.4–11.4; *P* = 0.01, respectively). Among incomplete the antibiotic regimen, there were less funisitis in the group with decreased *Ureaplasma* due to antibiotic therapy than in the group with increased or negative *Ureaplasma* at the time of PPROM.

**Fig 2.**
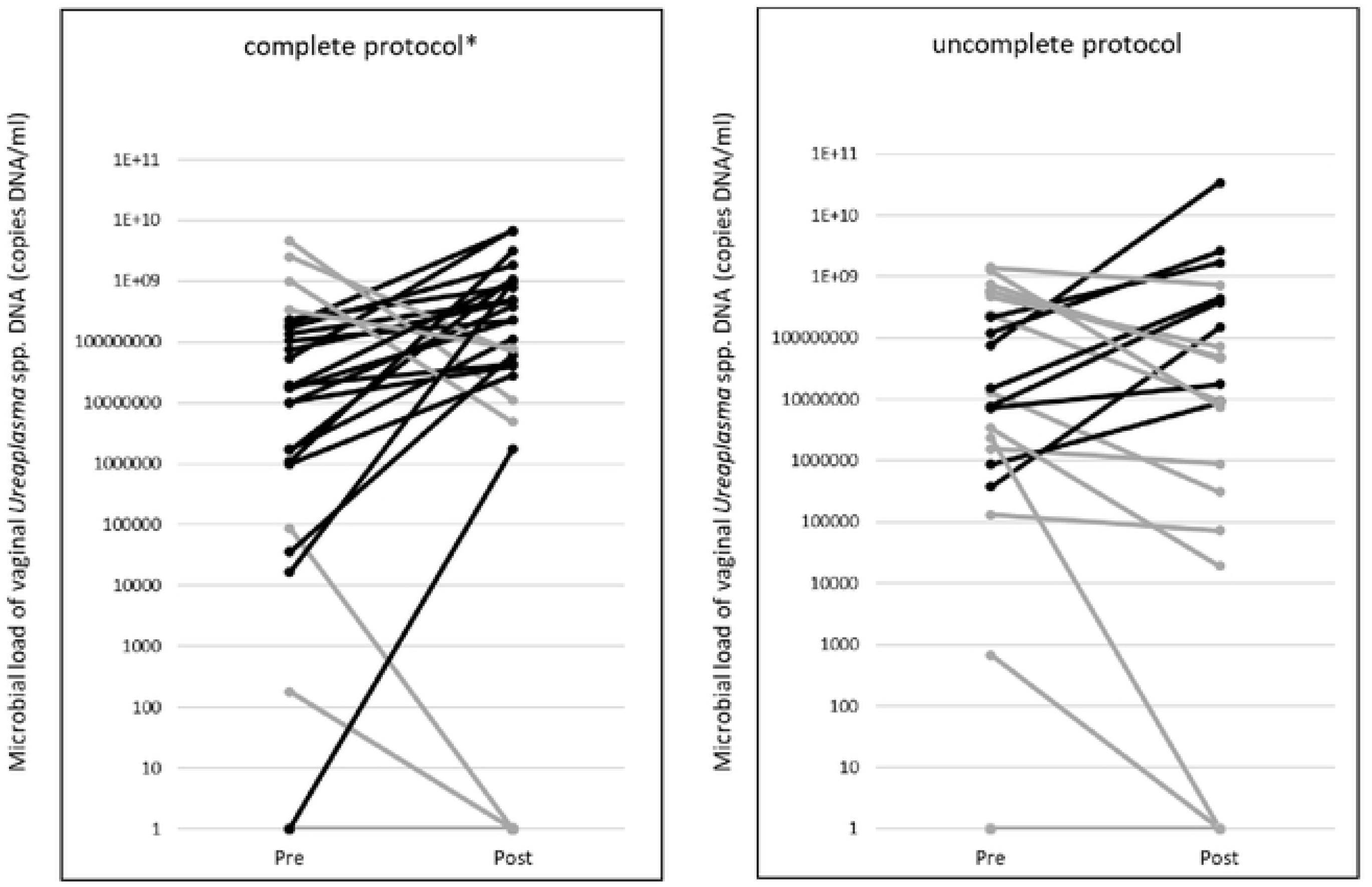
The change of microbial load of vaginal Ureaplasma DNA before and after antibiotic regimen. There were many cases of increased vaginal *Ureaplasma*, especially in patients who had completed the antibiotic regimen. * intravenous ampicillin 2 g every 6 hours for 2 days and a single oral dose of azithromycin 1 g followed by 5 days of oral amoxicillin 500 mg every 8 hours.

**Fig 3.**
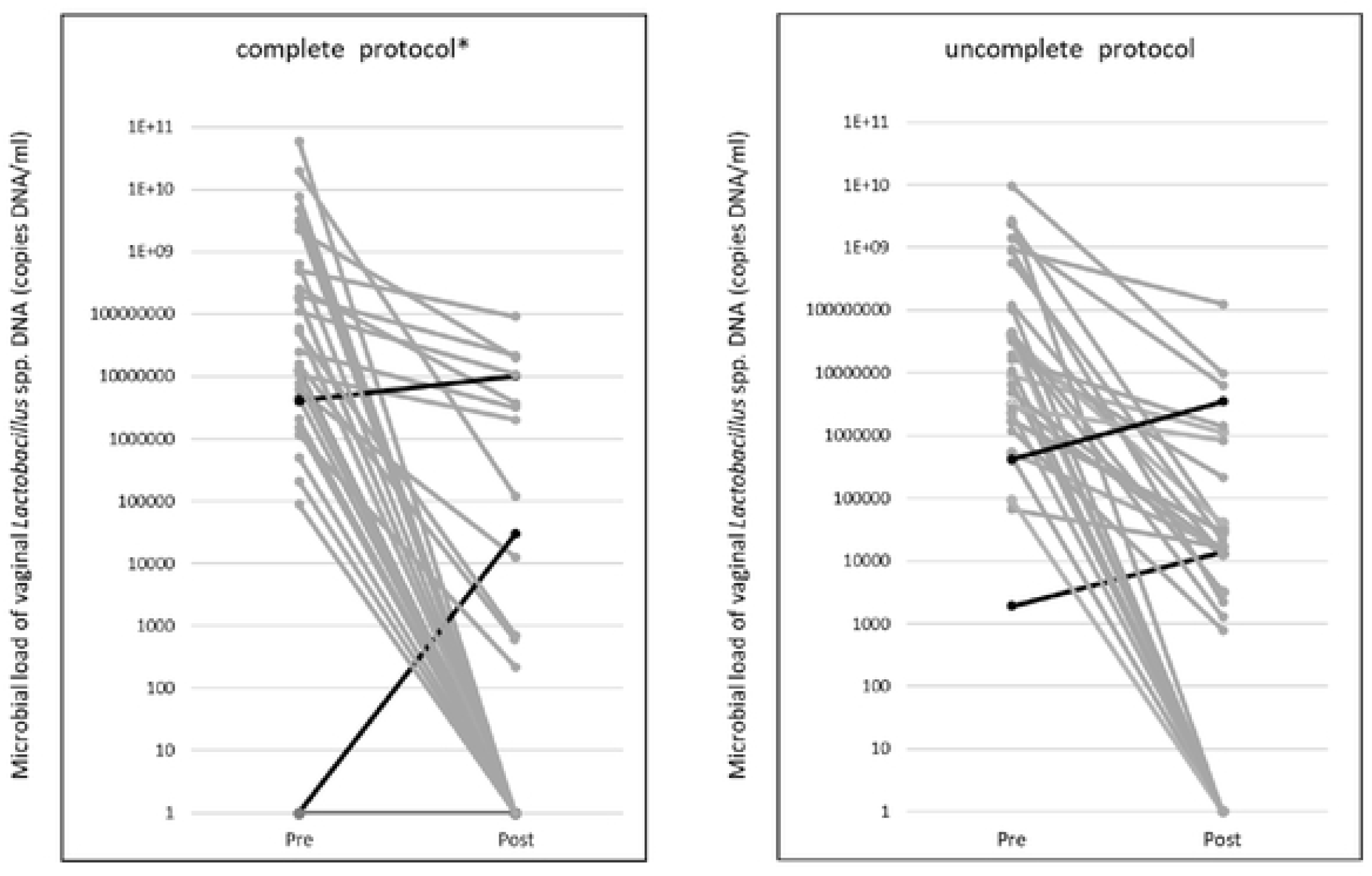
The change of microbial load of vaginal *Lactobacillus* spp. DNA before and after antibiotic regimen. Almost all *Lactobacillus* spp. were decreased or disappeared after the antibiotic regimen, especially in the cases that completed the antibiotic regimen. * intravenous 2 g ampicillin every 6 hours for 2 days, a single oral dose of 1 g azithromycin, and 5 days of oral 500 mg amoxicillin every 8 hours.

**Table 2.**
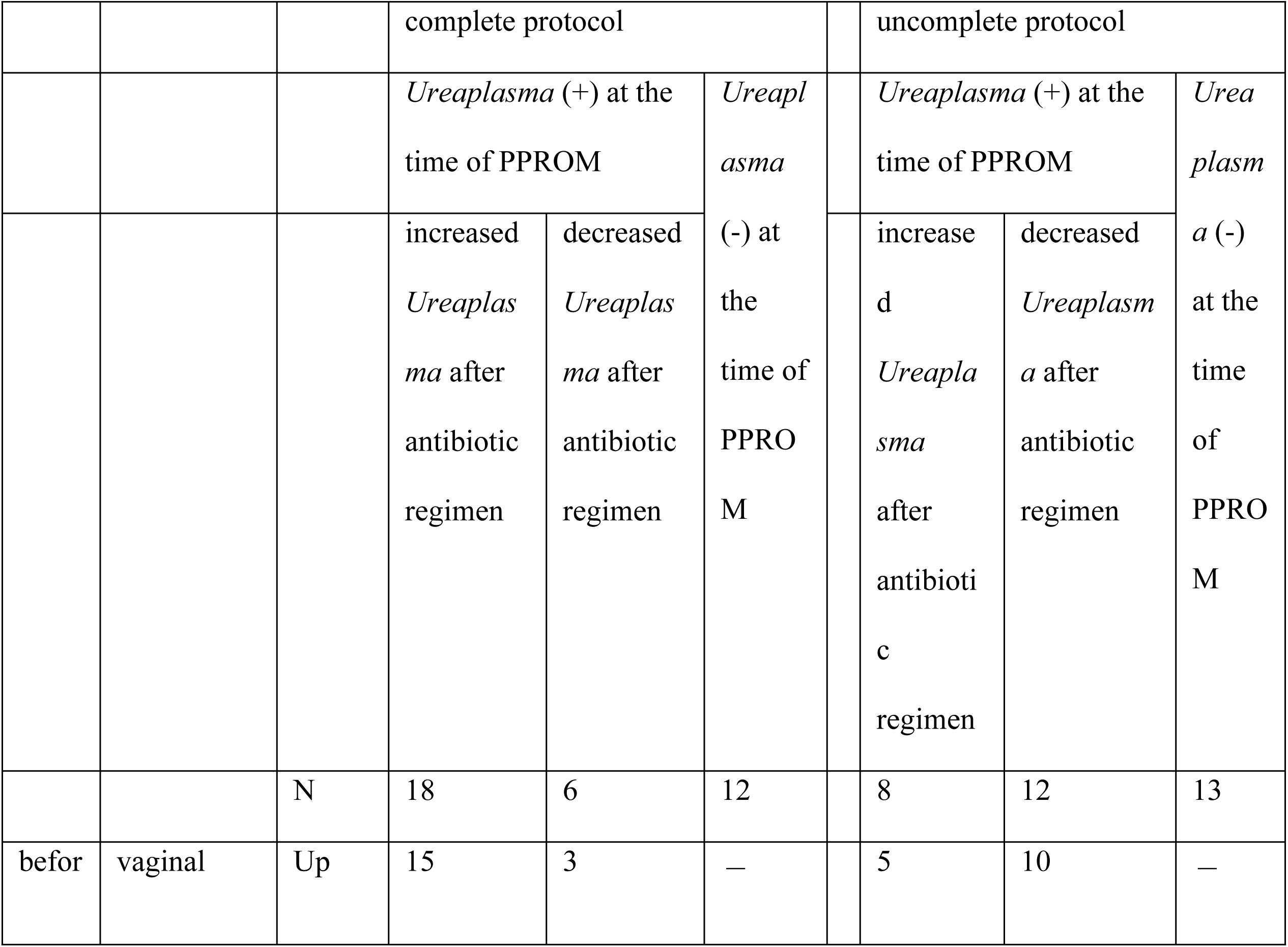
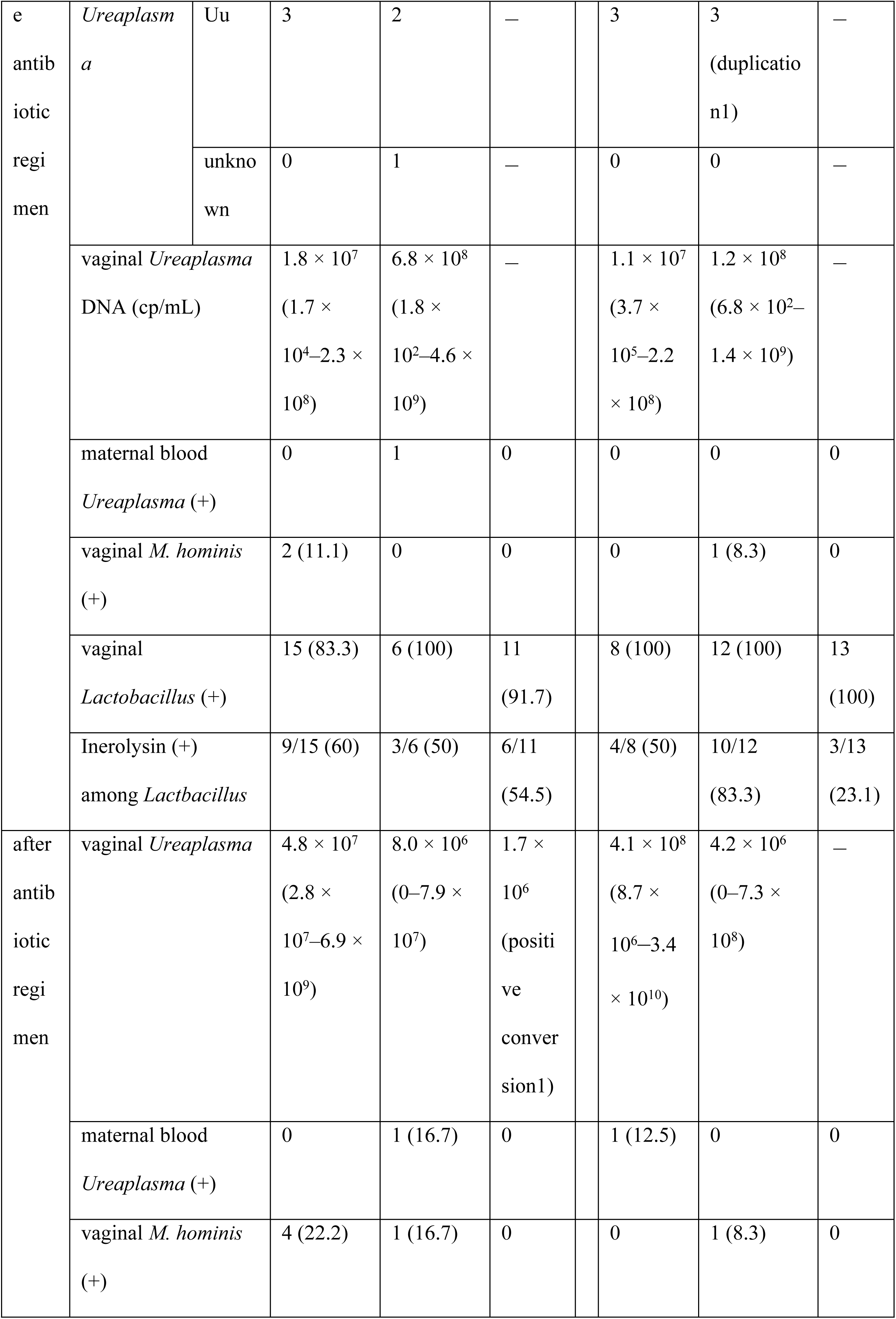
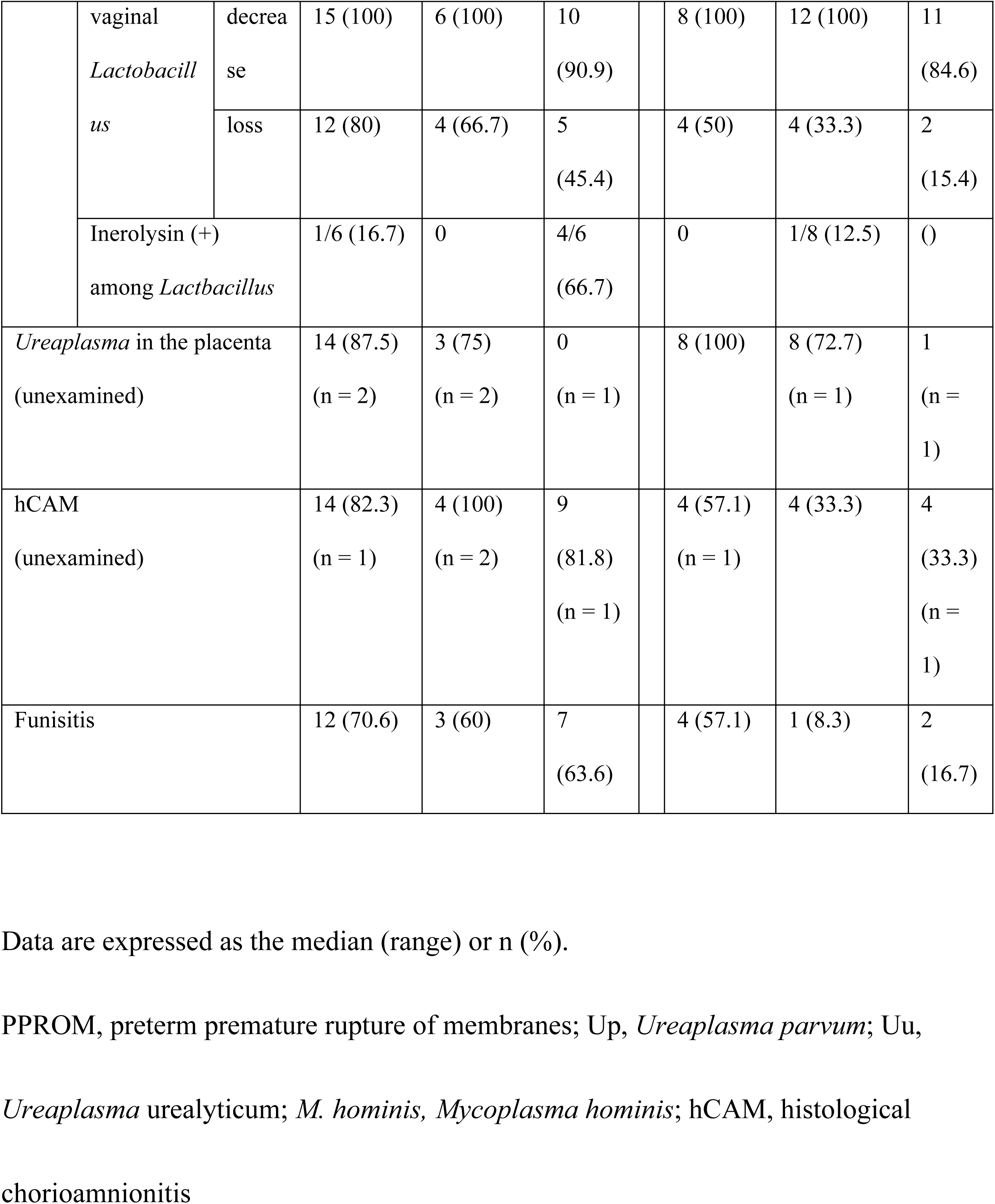
*Ureaplasma* in vagina and maternal blood, vaginal *Mycoplasma hominis*, vaginal *Lactobacillus spp*, and inerolysin classified by whether the antibiotic regimen was completed or not and how vaginal *Ureaplasma* was changed

Table 3 shows the neonatal complications according to vaginal *Ureaplasma* changes. Vaginal *Ureaplasma* changes were not associated with neonatal intensive care unit admission, neonatal sepsis, mechanical ventilation, or BPD. However, BPD was more frequent in those who completed protocol than in those who did not complete the protocol, regardless of vaginal *Ureaplasma* changes due to the antibiotic regimen, presence of *Ureaplasma* at the time of PPROM, and gestational age at delivery. (aOR, 7.2; 95%CI 1.3–40.1; *P =* 0.02)

**Table 3.**
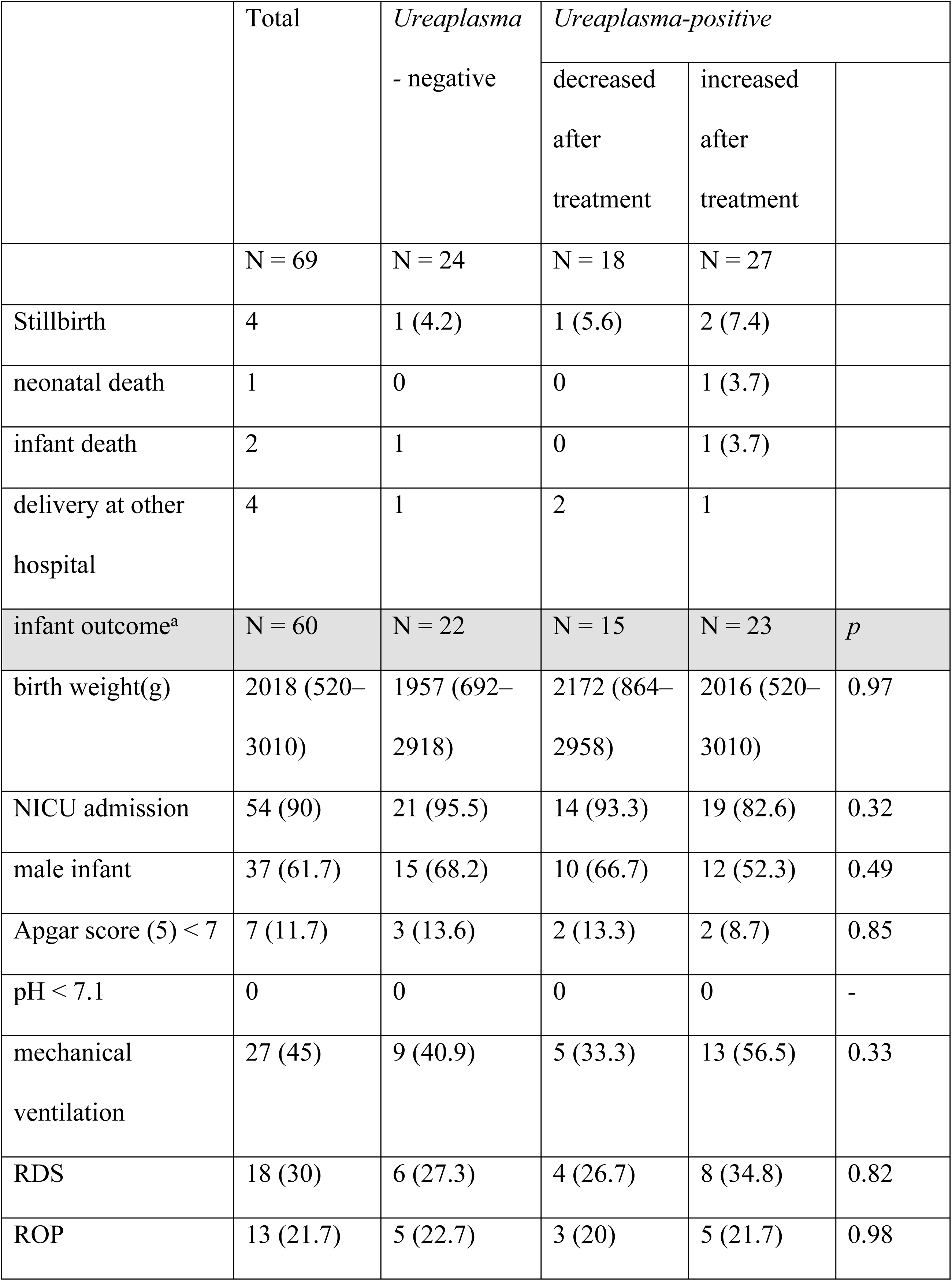
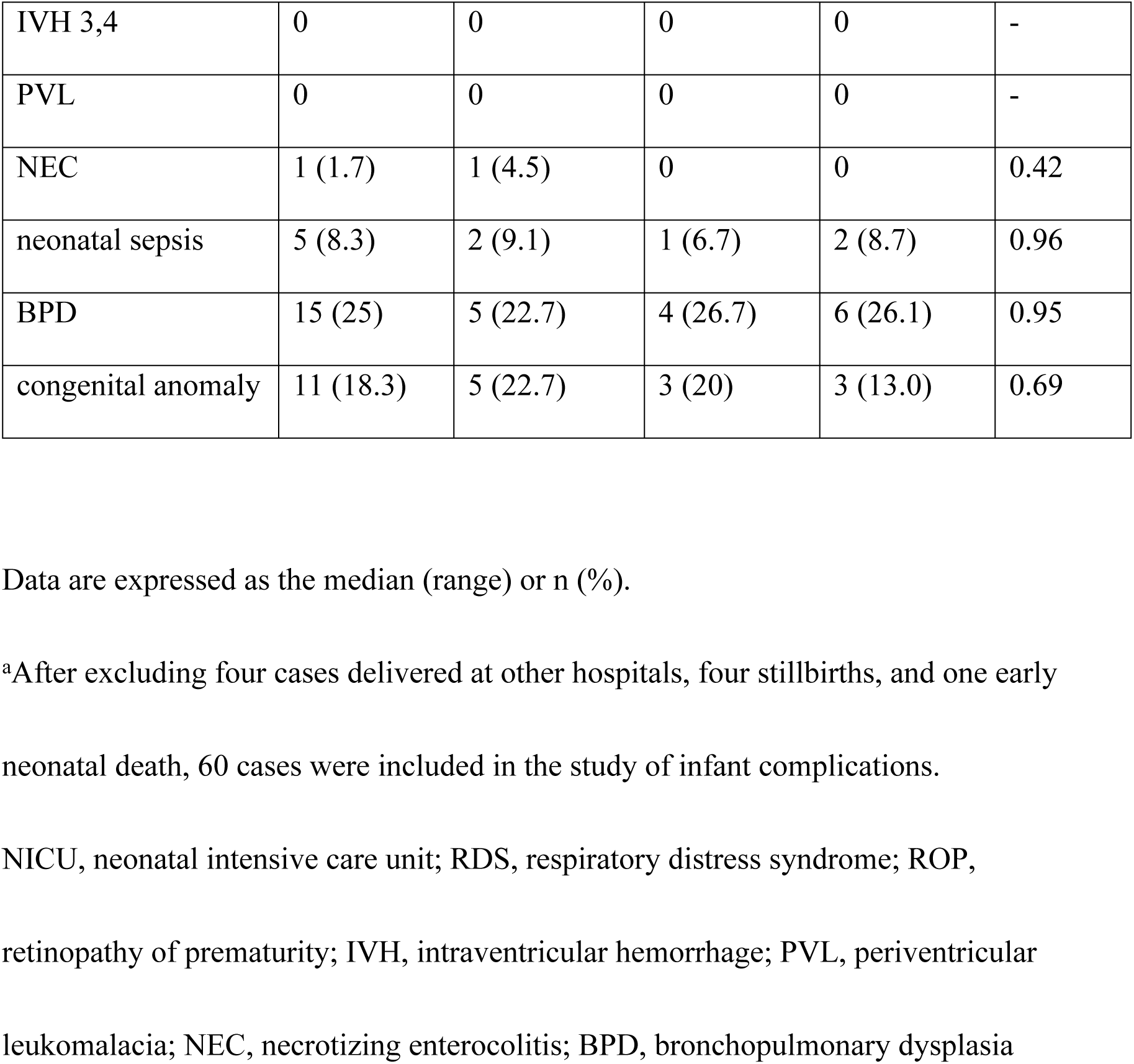
The neonatal complications according to vaginal *Ureaplasma* changes due to the antibiotic regimen.

## Discussion

Almost all *Lactobacillus* spp. decreased or disappeared, while most vaginal *Ureaplasma* and *M. hominis* increased after the antibiotic regimen, particularly in patients who completed the regimen. Patients with increased vaginal *Ureaplasma* had longer latency. This result did not indicate that cases of increased *Ureaplasma* would have a longer latency period. Rather, the longer period reflected *Ureaplasma* that was not reduced by antibiotic therapy grew intravaginally. The longer latency from PPROM may increase the likelihood of ascending infection of *Ureaplasma* to the amnion despite the antibiotic therapy. The possibility of avoiding invasive amniocentesis was considered if *Ureaplasma* could be detected in maternal blood in the presence of intrauterine *Ureaplasma* infection. However, it was impractical because *Ureaplasma* in maternal blood was undetectable in most cases.

Antibiotics were recommended for women with PPROM to prolong the delivery latency interval and reduce chorioamnionitis and neonatal sepsis.(8, 9, 17, 18) A regimen of intravenous ampicillin and erythromycin followed by oral amoxicillin and erythromycin has been commonly used. (8) Recently, azithromycin has been used instead of erythromycin because of its low cost, better side effect profile, ease of administration, and antibiotic resistance. The resistance of *Ureaplasma* and *Mycoplasma* to erythromycin was reported to be over 80%. (19) Azithromycin was associated with a similar latency interval and lower rate of clinical chorioamnionitis for women with PPROM.(12, 20) Tanaka reported the eradication of *Ureaplasm*a in the amniotic fluid of patients with PPROM treated with intravenous ampicillin–sulbactam and oral azithromycin.(21) Kacerovsky showed that intravenous clarithromycin was related to the reduced *Ureaplasma* DNA load in the amniotic fluid of patients with PPROM.(10) Conversely, Gomez reported that antibiotic administration (ceftriaxone, clindamycin, and erythromycin for 10–14 days) rarely eradicates intra-amniotic infection in patients with preterm PROM.(22) A report showed that azithromycin effectively eradicated respiratory tract *Ureaplasma* colonization in preterm infants. (23) In this study, most vaginal *Ureaplasma* was increased after the antibiotic regimen, suggesting an increasing resistance of *Ureaplasma* to azithromycin. In previous reports, the resistance rates of *U. urealyticum and M. hominis* to azithromycin also increased by about 14%–19.5%. (24, 25)

In this study, there was an increased frequency of hCAM, funisitis, and BPD in cases that completed the antibiotic regimen. An increased vaginal *Ureaplasma* increased the frequency of funisitis but not the incidence of neonatal infection and BPD. These suggest that even if the antibiotic regimen contributes to a prolonged gestational age, it may increase the frequency of intrauterine infection or neonatal infection from proliferated *Ureaplasma* and decreased *Lactobacillus* spp. Many studies presented that *Ureaplasma* was associated with adverse pregnancy outcomes and neonatal morbidities, such as preterm delivery, PPROM, chorioamnionitis, BPD,(6, 26) intraventricular hemorrhage,(27) and necrotizing enterocolitis.(28) Macrolides have anti-inflammatory properties that might reduce lung damage in preterm infants and treat bacterial infections(29); they reduce BPD in preterm infants. (30, 31) In this study, respiratory samples from infants were not obtained for *Ureaplasma* culture, so it was unknown whether the infants had *Ureaplasma* infection. The sample size was insufficient to validate the neonatal morbidity, so further research is needed.

*Lactobacillus*, the most frequently isolated microorganism from a healthy human vagina, is touted to prevent the invasion of pathogens and ensure vaginal epithelial homeostasis. (32) The loss of *Lactobacillus* and increase in Gram-negative rods, such as *Gardnerella vaginalis*, lead to bacterial vaginosis. (33) In a recent study*, L. crispatus*, *L. gasseri*, *L. jensenii*, and *L. iners*, were the most frequently isolated in the vaginas of healthy childbearing age or pregnant women. (34) A vaginal microbiota that is rich in *L. crispatus*, *L. gasseri*, and *L. jensenii* is often associated with a lower risk of bacterial vaginosis and preterm birth. Conversely, *L. iners* may play a role in the pathogenesis of bacterial vaginosis associated with preterm birth.(35, 36) *L. iners* alone detected in vaginal smears of healthy women in early pregnancy might be related to preterm delivery.(35) *L. iners* secretes the cholesterol-dependent cytolysins (CDC), inerolysin, as well as one of the CDC family, vaginolysin, produced by *G. vaginalis*. (37) In this study, about half of the cases had inerolysin, which may be related to the fact that the subjects in this study were cases of PPROM. There was no difference in the increase in vaginal *Ureaplasma* with or without inerolysin. *Lactobacillus* spp. was decreased after the antibiotics therapy regardless of the presence or absence of inerolysin. It is unclear whether *Lactobacillus* spp., reduced by antibiotic therapy, will recover spontaneously. It may be challenging to reduce *Ureaplasma* with this antibiotic therapy; we may rather focus on *Lactobacillus* formulations.

The strength of this study is that it is the first to investigate both vaginal *Ureaplasma* and *Lactobacillus* changes caused by ampicillin and azithromycin regimens. This was also an examination at a single institution with a uniform management policy. However, it has several limitations. In this study, vaginal cultures were collected rather than from amniotic fluid through transabdominal amniocentesis, which does not directly indicate amniotic fluid infection. There were three reasons why the specimens were vaginal secretions. First, the purpose of this study was to observe changes in both *Ureaplasma* and *Lactobacillus* due to antibiotic administration. Second, in PPROM cases, amniotic fluid testing after antibiotic administration might not be available due to the almost complete loss of amniotic fluid in the uterine cavity or the rapid onset of labor. Third, amniocentesis was invasive. In almost all vaginal *Ureaplasma-*positive cases, *Ureaplasma* was found on the surface of the placenta, regardless of the delivery mode and vaginal *Ureaplasma* changes due to the antibiotic regimen. Although there may be contaminations during delivery in PPROM cases, most vaginal *Ureaplasma* may be ascending into amniotic fluid or placenta. Another limitation is that the sample size was insufficient to validate the neonatal morbidity.

## Conclusions

Almost all *Lactobacillus* spp. decreased while most vaginal *Ureaplasma and M. hominis* increased after the antibiotic regimen. This suggests that *Ureaplasma* and *M. hominis* became resistant to azithromycin. Future studies are needed to revalidate current antibiotic therapy for PPROM because azithromycin-resistant *Ureaplasma* and *M. hominis* are widespread in the vaginal tract.

## Data Availability

All relevant data are within the manuscript and its Supporting Information files.

## Acknowledgements

We gratefully acknowledge the work of the medical staff of our center for their assistance in specimen collection.

